# Celebrex adjuvant therapy on COVID-19: An experimental study

**DOI:** 10.1101/2020.05.05.20077610

**Authors:** Wenxin Hong, Yan Chen, Kai You, Shenglin Tan, Feima Wu, Jiawang Tao, Xudan Chen, Jiaye Zhang, Yue Xiong, Fang Yuan, Zhen Yang, Tingting Chen, Xinwen Chen, Ping Peng, Qiang Tai, Jian Wang, Fuchun Zhang, Yinxiong Li

**Author notes:** These authors contributed equally to this work (Wenxin Hong, Yan Chen, Kai You, Shenglin Tan). Correspondence to: Dr Jian Wang, The Eighth People’s Hospital of Guangzhou, Guangzhou 510060, China.;, or, Prof Fuchun Zhang, The Eighth People’s Hospital of Guangzhou, Guangzhou 510060, China.;, or, Prof Yinxiong Li, Institute of Public Health, Guangzhou Institutes of Biomedicine and Health, Chinese Academy of Sciences, Guangzhou 510530, China.

## Abstract

**Background:** The world is under serious threat with the spread of the severe acute respiratory syndrome coronavirus 2 (SARS-CoV-2), which causes the coronavirus disease 2019 (COVID-19). However, there is no effective drug for the treatment of COVID-19. Based on analyses of available data, we deduced that the excessive prostaglandins E_2_ (PGE_2_) accumulation mediated by cyclooxygenase-2 (COX-2) was the key pathological basis of COVID-19.

**Methods:** The urine PGE_2_ levels were measured by mass spectrometry. An experimental study about Celebrex to treat COVID-19 was conducted based on routine treatment. A total of 44 confirmed COVID-19 patients were enrolled (Experimental group n=37, Control group n=7). Patients in experimental group were given Celebrex once or twice a day (0.2 g/time) for 7–14 days. The dosage or duration was modified for individuals. Clinical outcomes of Celebrex adjuvant therapy were evaluated by vital signs, laboratory tests, and computed tomography upon the discontinuance of Celebrex.

**Results:** We found that the concentrations of PGE_2_ in urine samples of COVID-19 patients were significantly higher than that of healthy individuals (mean value is 170 ng/ml *vs* 18.8 ng/ml, *p*<0.01) and positively correlated with the progression of COVID-19. Among the experimental group (ordinary n=29, severe n=7, critical n=1), 25 cases were treated with full dose and 11 cases with half dose of Celebrex, and 1 case with Ibuprofen. The remission rate were 100%, 82% and 57% in full dose, half dose and control group respectively. Celebrex significantly reduced the PGE_2_ levels and promoted recovery of ordinary or severe COVID-19.

**Conclusion:** Our study suggests that Celebrex adjuvant treatment may be helpful for the therapy of COVID-19.

## Introduction

The severe acute respiratory distress syndrome (ARDS) caused by SARS-CoV, MERS-CoV and SARS-CoV-2 infections is a major factor of mortality. It has been found that the nucleocapsid protein (N) and spike glycoprotein (S) of SARS-CoV can directly bind to the promoter of cyclooxygenase-2 (COX-2) gene, which drives overexpression of COX-2 in a dose-dependent manner.[1, 2] Homologous analysis showed that there were 90.6% and 75.8% similarity of the N and S protein between SARS-CoV and SARS-CoV-2. Therefore, it is possible that the SARS-CoV-2 infection might also hold the potential to induce COX-2 overexpression in lung epithelial cells, resulting in a significant accumulation of prostaglandins, especially prostaglandin E_2_ (PGE_2_).

Excessive PGE_2_ levels may participate in COVID-19 pathology with one of the following mechanisms: 1) binding to the EP2 receptor causing fever, pain, acute inflammation, and enhanced vascular permeability; 2) binding to the EP3 receptor leading to edema, inflammatory mucus secretion, increased viscosity of exudates covered alveoli, bronchioles blockage, thus hindering blood oxygen exchange;[3] 3) binding to the EP4 receptor, causing bronchial contractions and spams,[4] increasing airway resistance, causing respiratory and hemodynamic disorders, ARDS and multi-organ failures; 4) inhibition of T lymphocyte functionality, by promoting amplification, differentiation and proliferation of Th1 and Th17 subtypes through EP4 receptor causing contact hypersensitivity of bronchioles;[5] 5) PGE_2_ and thromboxane A2 (TXA2) activate platelet aggregation and thrombosis, contributing to pulmonary hypertension in ischemia-reperfusion lung injury.[6]

Under the circumstances of this international emergency and the situation of having no effective drugs for COVID-19, it makes great sense to explore the pathological mechanisms of COVID-19 and to establish an integrated strategy for diagnosis and treatment with available drugs using the discovered key pathological target(s).

Here, we propose that excessive PGE_2_ may be a key in the pathology of COVID-19 and that COX-2 is the critical target for therapy. To test this hypothesis, the urinary PGE_2_ levels were determined in COVID-19 patients to verify its correlation with disease status. And Celebrex, a specific inhibitor of COX-2, was to be used for experimental study.

## Methods

### Study design and participants

This was a prospective study done at Guangzhou Eighth People’s Hospital. Patients with SARS-CoV-2 infection were confirmed by next-generation sequencing or real-time RT-PCR according to a previously published protocol.[7] Based on the “Diagnosis and Treatment Guideline for COVID-19” of China, the clinical criteria for classification of OVID-19 stage in Chinese Guideline are listed as following. Mild: The clinical symptoms are mild and no pneumonia manifestation can be found in CT imaging; Ordinary: Fever and respiratory tract symptoms, etc. and pneumonia manifestation can be seen in CT imaging; Severe: Meeting any of the following: 1) Respiratory distress, RR≥30 breaths/min; 2) Oxygen saturation ≤ 93% at a rest state; 3) Arterial partial pressure of oxygen (PaO2)/oxygen concentration (FiO2) ≤ 300mmHg; Critical: Meeting any of the following: 1) Respiratory failure occurs and mechanical ventilation is required; 2) Shock occurs; 3) Complicated with other organ failure that requires Intensive care unit (ICU) care.

A total of 44 confirmed COVID-19 patients, who were admitted to hospital from January 23 to February 15, 2020, were enrolled into this study (Table 1 and Table S1). One of the 44 patients were classified as critical case (2.2%), 7 of the 44 patients were classified as severe type cases (16%) and the others were ordinary type cases (81.8%). The enrolled patients are fully aware of the purpose, benefits and potential risks, and signed the informed consent prior to this study. This investigational study design was approved by the Medical Ethics Committee of Guangzhou Eighth People’s Hospital (Approve number: AF/sc-02/01.6). The study was registered on the Chinese Clinical Trials Registry, ChiCTR2000031630.

**Table 1.** Characteristics and clinical outcomes of COVID-19 patients^*^

| **Table 1. Characteristics and clinical outcomes of COVID-19 patients \*** |  |  |  |  |
| --- | --- | --- | --- | --- |
| **Parameters** | **All patient**<br>**(n=43)** | **Celebrex group (Oral 0.2 g)** |  | **Control group**<br>**(n=7)** |
|  |  | **BID (n=25)** | **QD (n=11)** |  |
| **Characteristics** |  |  |  |  |
| Age, years | 49.5 (15.3) | 45.8 (12.8) | 57.2 (16.6) | 49.5 (18.3) |
| 0-14 | 0 | 0 | 0 | 0 |
| 15-44 | 18 (42%) | 12 (48%) | 2 (18%) | 4 (57%) |
| 45-64 | 18 (42%) | 12 (48%) | 5 (46%) | 1 (14%) |
| ≥65 | 7 (16%) | 1 (4%) | 4 (36%) | 2 (29%) |
| Female sex | 22 (51%) | 14 (56%) | 5 (46%) | 3 (43%) |
| **Clinical outcomes after discontinuation of Celebrex** |  |  |  |  |
| Remission | 38 (88%) | 25 (100%) | 9 (82%) | 4 (57%) |
| Exacerbation | 5 (12%) | 0 | 2 (18%) | 3 (43%) |

\* Data are mean (SD) or n (%); Reduced denominators indicate missing data. Percentages may not total 100 because of rounding. BID twice a day; QD once a day.

### Clinical information and Celebrex usage

The clinical information including the physical, laboratory tests and chest computed tomography (CT) of COVID-19 patients were collected.

The patients in experimental group were treated with Celebrex (Celecoxib, Pfizer, Dalian, China) in combination with routine treatments suggested by the guideline. The usage and dose of Celebrex was once or twice a day (0.2 g/time) for 7–14 days by oral. The dosage or duration of medication was subject to change based on each individual case. Routine treatments were according to the National Guideline, including isolation, nursing, bed rest, symptomatic and supportive treatment, antibiotics, antiviral medication, glucocorticoids, oxygen therapy and/or assisted breathing.

Based on the comparison of sequential chest CT images, as well as the changes of symptoms and laboratory test results, the clinical outcomes will be classified with three categories included remission, constant and exacerbation. Remission was defined as dissipating/clarifying of the mass opacities in chest CT images, decreasing the D-dimer, CRP, serum alanine aminotransferase and aspartate aminotransferase levels and improvement of lymphopenia and neutrophilia; the constant and exacerbation outcomes were evaluated accordingly with the changes of those parameters. The length of Celebrex treatment was determinate by the individual conditions. The discharge standards according to the national guideline are listed as following: 1) with normal body temperature for more than 3 days; 2) with significantly recovered respiratory symptoms; 3) lung imaging shows obvious absorption and recovery of acute exudative lesion; 4) with negative results of the nucleic acid tests of respiratory pathogens for consecutive two times (sampling interval at least 1 day).

### The measurement of urine PGE_2_ by mass spectrometry

Agilent 1290 Infinity II High Performance Liquid Chromatograph (HPLC) was used in conjunction with Agilent 6470 Triple Quadrupole Mass Spectrometer for the PGE_2_ measurement with modifications to the previous method.[8] The sample was mixed with 3x volume of ethanol to inactivate the virus and the resulted supernatant was mixed with an equal volume of methanol. After being centrifuged at 13000*g* for 15 minutes, the supernatant was mixed with 2x volumes of ddH_2_O (0.1% HCOOH) and analyzed by the Agilent LC-QQQ 6470 with a 1290 HPLC. PGE_2_ was separated by using Zorbax Eclipse plus C18 3.0×150 mm (1.8 μm) with a 10 minute linear gradient acetonitrile (0.1% HCOOH), and measured by monitoring *m/z* 351.1 to 271.3 under MRM with negative ion.

### Statistical analysis

The differences between two groups of data were compared by Student’s-*t* test, and the statistical results were expressed by mean ± standard error (mean ± SEM). The statistical analysis was conducted by using GraphPad Prism 7 software. *p* <0.05 was considered statistically significant.

## Results

The concentrations of PGE_2_ in urine samples were determined by a method of mass spectrometry (Figure S1). Our data showed that the PGE_2_ levels in COVID-19 patients, who were hospitalized within two days, were significantly higher than the ones of healthy individuals (170±40 ng/ml *vs* 18.8±3.8 ng/ml, *p*<0.01) (Figure 1). We determined that the normal threshold of PGE_2_ concentration in urine is lower than 20 ng/ml and that 100 ng/ml is considered to be significant as a risk line.

**Figure 1:**
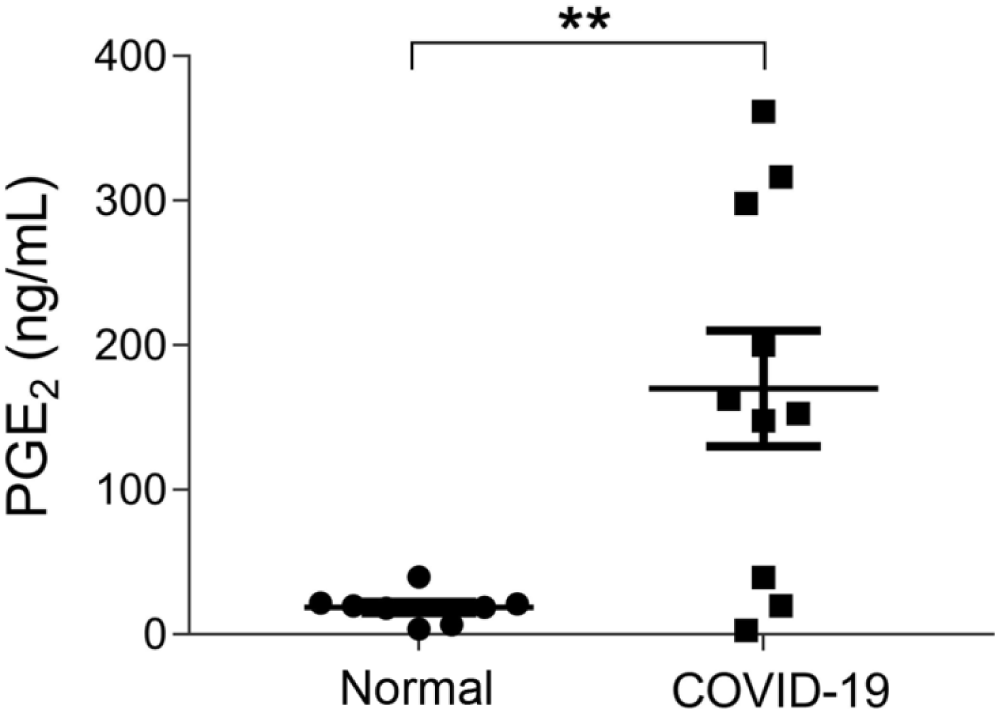
The urinary PGE_2_ levels of COVID-19 patients were significantly increased. The urinary PGE_2_ concentrations of COVID-19 patients, who were hospitalized within 2 days, were significantly higher than the health individuals (170±40 ng/ml *vs* 18.8±3.8 ng/ml, *p*<0.01). **: *p* < 0.01.

Since the PGE_2_ was mainly generated by COX-2, then a COX-2 specific inhibitor (Celebrex) was used to treat COVID-19 patients based on the routine treatment. On March 19, 2020, 25 cases (6 severe, 19 ordinary) were given full dose (0.2g, twice a day) of Celebrex, and all cases showed an improved outcomes after discontinuation. There were 11 cases (2 severe/critical, 9 ordinary) that had received half dose (0.2g, once a day) of Celebrex, and all the ordinary cases showed improvement, while the 2 severe/critical cases were exacerbated after discontinuation of treatment. Whereas in control group (n=7), 4 cases were improved and 3 cases were exacerbated at day 10 after admission in hospital (Table 1 and Table S1). Our results indicated that Celebrex treatment with a conventional dose (0.2 g, twice a day) might effectively promote the recovery of ordinary and severe cases of COVID-19.

It has been reported that 15.7% of the ordinary COVID-19 cases progressed to a severe stage under routine treatments.[9] With Celebrex treatment, none of the 29 ordinary cases progressed to a severe classification (Table S1). Two cases were chosen to represent the control (case C1) and experimental group (case E20) respectively (Figure 2). The PGE_2_ level of C1 remained at a high level (>1000 ng/ml) during day 5–9 (Figure 2A). While the case E20 treated with Celebrex decreased steadily from over 1000 ng/ml (day 2) to 100 ng/ml (day 3–4) (Figure 2B). Except the improvements of symptoms and laboratory tests, the chest CT images (at day 4–10) showed that the improvement of pulmonary opacification and pneumonia of case E20 was faster than the control group C1 case in the same period (Figure 2C and D). It suggested that Celebrex might promote the recovery process of ordinary COVID-19 patients.

**Figure 2:**
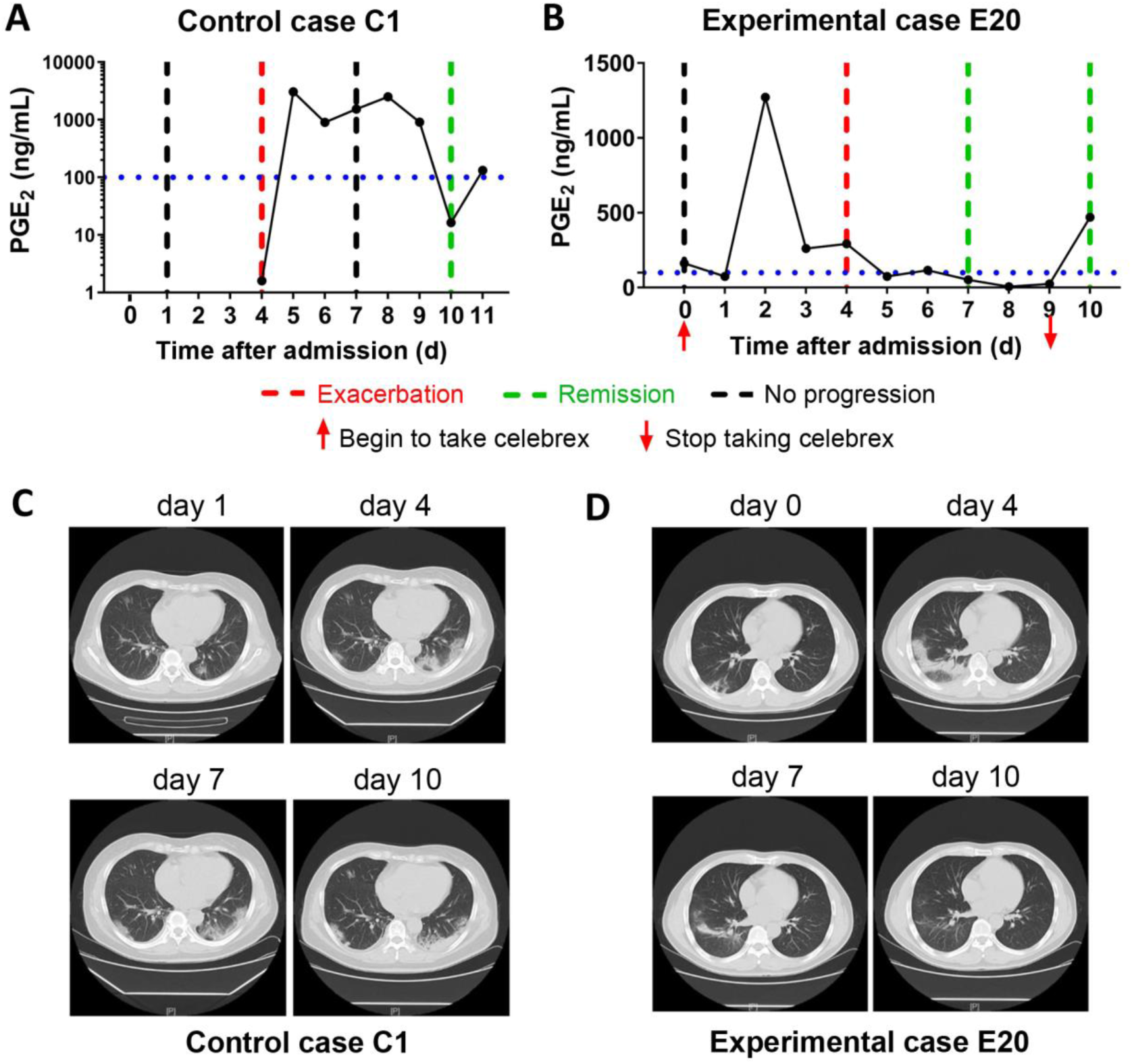
Celebrex treatment accelerated the recovery of those ordinary COVID-19 patients and prevented the progression towards severe stage. A: Control case C1, the dynamic changes of the urinary PGE_2_ were correlated with the COVID-19 prognosis; B: Experimental case E20, the dynamic reduction of PGE_2_ were matched with the improvements of COVID-19 conditions; C and D: The representation of sequential chest CT images illustrated therapeutic outcomes of these two cases respectively.

In addition, none of the severe COVID-19 cases with full dose of Celebrex treatment progressed to critical illness. The comparison of the control (case C2) and experimental cases (case E6) with similar severe pulmonary opacification diagnosed by CT imaging was illustrated (Figure 3). The PGE_2_ levels of case C2 fluctuated around 1000 ng/ml at day 5-12 after admitted to hospital with routine treatment (Figure 3A). In contrast, the PGE_2_ levels of case E6 in Celebrex group were steady decrease, and remained at lower level than 100 ng/ml (Figure 3B). Indeed, the chest CT images (at day 3, 6 and 11) of case E6 showed that the ground glass-like opacities were clarified continually, and significantly faster than the control case C2 (Figure 3C and D). It suggests that Celebrex might reverse the progress of severe COVID-19 and prevented the progression to a critical stage.

**Figure 3:**
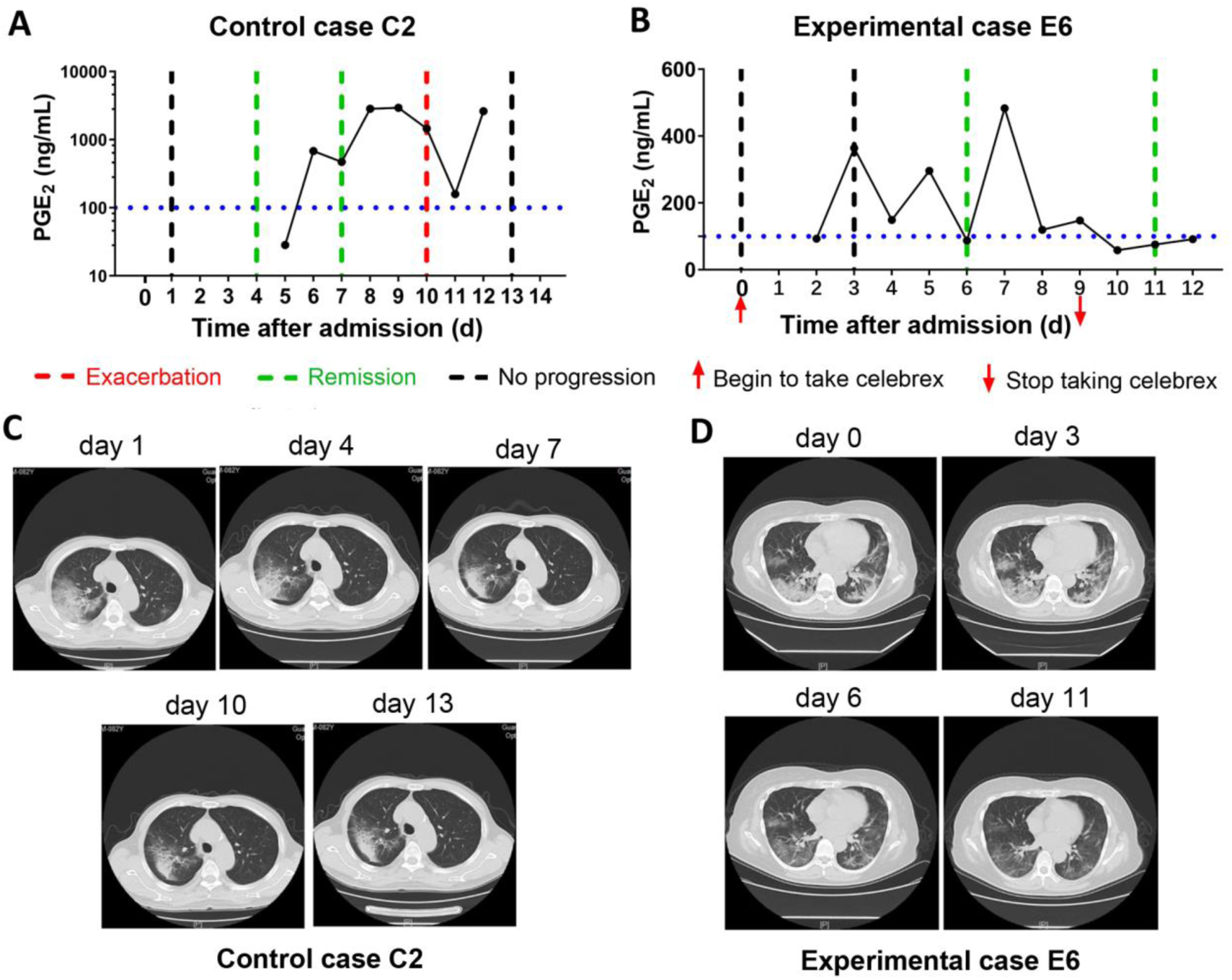
Celebrex treatment promoted the improvement of severe stage of COVID-19 and blocked the progression towards critical stage. A: Control case C2, the dynamic changes of the urinary PGE_2_ were correlated with its COVID-19 prognosis; B: Experimental case E6, the dynamic reduction of PGE_2_ were matched with the improvements of COVID-19 conditions; C and D: The representation of sequential chest CT images illustrated therapeutic outcomes of these two cases respectively.

Moreover, there were two patients (experimental case E3 and E5), who were hospitalized and received routine treatment for 12 and 15 days respectively, progressed from ordinary to severe illness. After taken Celebrex based on routine treatment, their PGE_2_ levels were controlled and the pneumonia were gradually improved (Figure 4). These findings indicated that Celebrex may also be effective on patients who progressed from ordinary into severe type under routine therapy.

**Figure 4.**
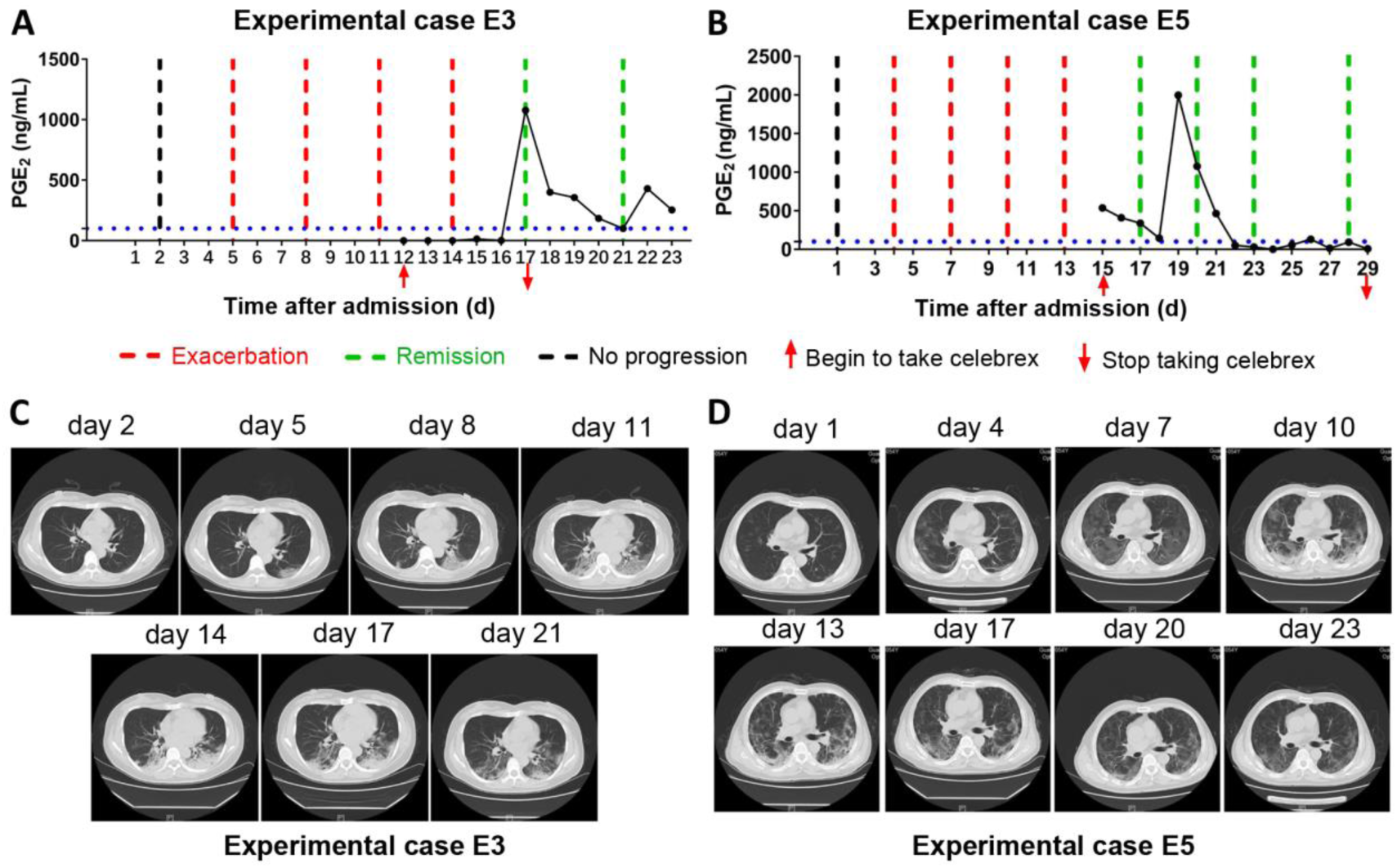
Celebrex intervention reversed the progressed severe stage under routine treatment. A and B: Two ordinary cases were received routine treatment for 12 days and 15 days respectively, and the conditions gradually progressed into severe stage, upon the Celebrex treatment, the PGE_2_ levels were decreased along with the improvements of COVID-19; C and D: The representation of sequential chest CT images illustrated therapeutic outcomes of these two cases under Celebrex intervention respectively.

Altogether, our findings suggested that the increase of PGE_2_ may be critical for progression of COVID-19 and Celebrex intervention may effectively promote the recovery of ordinary and severe types of COVID-19.

## Discussion

ARDS is the leading cause of death of the critical SARS-CoV, MERS-CoV and SARS-CoV-2 infected patients. Autopsy reports indicate that the pathological manifestation of lung injury were diffuse alveolar injuries, including fibrin mucus exudation, ground-glass edema membrane formation, and alveolar epithelial cell detachment.[10–12] However, the pulmonary fibrosis and consolidation caused by SARS-CoV-2 was less severe than SARS-CoV, but the alveolar mucous secretion was more severe.[13] Therefore, it was speculated that the COVID-19 patient’s alveolar blood and gas exchange was seriously blocked, which led to ARDS. The patient eventually died from respiratory and multi-organ failures.

Prostaglandin is a class of small lipid molecules transformed from arachidonic acid by COX-1 and COX-2, and PGE_2_ is one of the most active molecule.[14] The COX-2/PGE_2_ pathway plays a crucial role in mucus secretion, desensitization of the β-2 adrenergic receptor, and the matrix metalloproteinase-mediated airway remodeling, cough, fever, asthma and other respiratory diseases.[15] It has been reported that both the N and S proteins of SARS-CoV could induce the expression of COX-2 in epithelial cells.[1, 2] The similarity of N and S protein sequences of SARS-CoV-2 and SARS-CoV were 90.6% and 75.8% respectively. It was found that both viruses infected host cells by using the S protein to bind Angiotensin I Converting Enzyme 2 (ACE2) receptor.[16] Thus, It suggests that SARS-CoV-2 might also have the ability to induce the expression of COX-2 in lung epithelial cells.

We confirmed that the urine PGE_2_ concentration of COVID-19 patients was significantly higher than healthy individuals. Celebrex, a specific inhibitor of COX-2, effectively decreased the level of urinary PGE_2_ and promoted the recovery of COVID-19. However, we also found that, in some cases, discontinuation of Celebrex might lead to increased PGE_2_ and pneumonia relapse, such as experimental cases E1 and E9, whose PGE_2_ levels were rebounded and increased quickly accompanied with slightly worsen of pulmonary opacification (Figure S2). These observations suggested that the duration of Celebrex treatment should be determined according to each patient’s condition to reduce the risk of disease relapse, deterioration and pulmonary fibrosis.

The half-life of PGE_2_ *in vivo* is about one minute.[17] After going through the lung, liver and kidney organs via blood circulation, up to 90% of PGE_2_ will be degraded. Extremely high concentrations of PGE_2_ (up to 2500 ng/ml) were detected in the urine of COVID-19 patients, such as control case C1 and C2 (Figure 2 and 3), so it is conceivable that the PGE_2_ levels in lung tissues and blood could be much higher. This kind of “prostaglandin storm” occurs in the very early and progressive stages of COVID-19. While very few immune cytokines could be detected in blood or urine during this early period, which could be occurred at the critical stage.

Currently, Zheng *et al* reported that in an influenza virus-infected mouse model of H5N1, the survival rate of mice treated with zanamivir alone (antiviral drug) was 13.3%. While combining zanamivir with Celebrex (COX-2 inhibitor) or with mesalazine (COX inhibitor) could improve the survival to about 20%. However, a combination of these three drugs significantly reduced viral load and increased survival up to 53.3%.[18] Although there is no specific drug for the treatment of COVID-19 today, we supposed that Celebrex combined with antivirus and other anti-inflammatory drugs treatment might be a good strategy for COVID-19 treatment.

There are no reports on whether Celebrex is involved in inhibiting SARS-CoV-2 replication. Although some studies have found that COX-1 and COX-2 inhibitors have the potential to inhibit replication of other subtypes of coronavirus. For example, Raaben *et al* reported that indomethacin and curcumin (COX non-selective inhibitors) inhibited the synthesis of RNA, protein, and production of virus particles of mouse hepatitis coronavirus in a dose-dependent manner. *In vitro* experiments showed that both SC-560 (COX-1 inhibitor) and NS-398 (COX-2 inhibitor) reduced mouse hepatitis virus (coronavirus) infection by 65–75% at concentrations that were nontoxic to the cells.[19] Santoro *et al* reported that indomethacin directly inhibited the production of SARS-CoV and CCoV virus particles, reduced cell infection *in vitro* and *in vivo*.[20]

Taken together, COX-2 overexpression accompanied with PGE_2_ accumulation may be a key in the molecular pathology of COVID-19. Celebrex, a specific COX-2 inhibitor, may be an effective drug for the treatment on COVID-19. However, our study was not a rigorous randomized, double-blind and controlled clinical trial. There is needed another well-designed large-scale clinical trial to validate this hypothesis. Our study may provide useful information for the treatment of COVID-19.

## Data Availability

The datasets used and/or analyzed during this study are available from the corresponding author on reasonable request.

## Ethics approval and consent to participate

This study design was approved by the Medical Ethics Committee of Guangzhou Eighth People’s Hospital.

## Consent for publication

Not applicable

## Contributors

JW, FZ, and YL conceived and designed the study. YC, KY, ST, and FW contributed to the literature search. WH, YC, KY, ST, FW, JT, XuC, JZ, YX, YF, ZY, TC, PP and QT contributed to data collection. WH, YC, KY, ST, FW, and JT contributed to data analysis. WH, XuC, XiC, and PP contributed to data interpretation. WH, YC, KY, ST, and JT contributed to the figures. YC, KY, ST, FW, JT, and YL contributed to writing of the report.

## Acknowledgements

This project was conducted in the national emergency period; we would like to thank the Guangdong General Police Hospital, Guangdong He Yi Company, Shenzhen Giant Future Company and Guangzhou Zhongke Baier Company for donating protective uniforms, masks, bio-safe transport equipment and other materials, as well as the many volunteers for their selfless dedication and hard work.

